# Increased subpial cortical lesion detection at 3 tesla using Inversion Recovery Susceptibility Weighted Imaging with Enhanced T2 Weighting (IR-SWIET)

**DOI:** 10.64898/2026.08.07.26359605

**Authors:** Edward Sizer, Kamso Onyemeh, Amit Kohli, Elle Levit, Chantal Roy-Hewitson, Zoe Brown, Jane Low, Ken Feb, Jiming Zhang, Adam Ulano, Francesco La Rosa, Govind Nair, Daniel S. Reich, Russell T. Shinohara, Sarah A. Morrow, Andrew J. Solomon, Erin S. Beck

## Abstract

**Background:** Multiple sclerosis subpial cortical lesions are prevalent and associated with disability but difficult to detect on MRI. Inversion recovery susceptibility weighted imaging with enhanced T_2_ weighting (IR-SWIET) and T_1_/T_2_ ratio imaging have been proposed for cortical lesion detection on 3 tesla (T) MRI.

**Objectives:** To assess cortical lesion detection using IR-SWIET and T_1_/T_2_ ratio imaging.

**Methods:** Cortical lesions were identified in 20 persons with MS (pwMS) independently on six image sets: T_1_ weighted (w) magnetization prepared 2 rapid acquisition gradient echoes (MP2RAGE) + T_2_w fluid attenuated inversion recovery (FLAIR) alone or with T_1_/T_2_, IR-SWIET single acquisition (×1), average of two (×2) or median of four (×4) acquisitions, or denoised single acquisition (IR-SWIET×1DN). In 10 additional pwMS with 7T-based cortical lesion segmentations, lesions were identified on MP2RAGE + FLAIR + IR-SWIET×1DN.

**Results:** Median subpial lesions identified on MP2RAGE + FLAIR was 0 (interquartile range (IQR) 2) vs 0 with T_1_/T_2_ (IQR 1, p=0.07), 1 with IR-SWIET×1 (IQR 6, p=0.42), 5 with IR-SWIET×2 (IQR 5, p=0.008), 4 with IR-SWIET×4 (IQR 6, p=0.008), and 4 with IR-SWIET×1DN (IQR 6, p=0.008). Versus 7T, IR-SWIET×1DN detected subpial lesions with similar sensitivity to IR-SWIET×2.

**Conclusions:** IR-SWIET, but not T_1_/T_2_, improves subpial cortical lesion detection. Denoising may be an efficient and sensitive alternative to multi-acquisition averaging.

## Introduction

Multiple sclerosis (MS) cortical lesions are commonly seen on histopathology and ultra-high field (7 tesla, T) MRI.^1–4^ Despite inclusion of cortical lesions in the 2017 and 2024 McDonald diagnostic criteria, visualization of cortical lesions on 3T MRI remains challenging.^5,6^ The most commonly used MRI sequence at 3T for cortical lesion detection, double inversion recovery (DIR), demonstrates good sensitivity for leukocortical lesions but only 7% sensitivity for subpial lesions.^7^ T_1_ weighted (w) magnetization prepared 2 rapid acquisition gradient echoes (MP2RAGE) achieves a similar cortical lesion detection rate with better grey-white matter contrast, but likewise is insensitive to subpial lesions.^8,9^ Subpial lesions are of particular interest as they are the most frequent subtype of cortical lesion in MS, are associated with disability and progressive disease, may form independently from white matter lesions and other types of cortical lesions, and might be specific to MS.^1,3^

We recently developed inversion recovery susceptibility weighted imaging with enhanced T_2_ weighting (IR-SWIET), an MRI method designed to improve subpial lesion detection at 3T by increasing T_2_ and T_2_* weighting and suppressing cerebrospinal fluid signal.^9^ In a single center study including ten persons with MS (pwMS) with known cortical lesions on 7T imaging, 3T IR-SWIET improved subpial lesion detection compared to other 3T methods, including DIR.^9^

At the sub-millimeter resolution required to image cortical lesions, IR-SWIET images are limited by low signal-to-noise ratio (SNR). Averaging multiple IR-SWIET acquisitions enhances SNR, lesion-to-background contrast, and subpial lesion detection, at a substantial cost in scan time.^9^

An alternative is a post-acquisition denoising algorithm such as non-local means filtering.^10, 11^

The sensitivity of denoised IR-SWIET images for cortical lesions has not previously been tested.

Another method that has been proposed to increase cortical lesion conspicuity at 3T is T_1_/T_2_ ratio images, in which T_1_ and T_2_w images are co-registered and divided.^12,13^ Contrast between cortical lesions and normal-appearing cortex is greater on T_1_/T_2_ images than on T_1_ or T_2_w images alone or on IR-SWIET, and lesion conspicuity is improved relative to conventional sequences in clinical trial data but prospective cortical lesion detection on T_1_/T_2_ images has not been tested.^12,13^

The primary aim of this study was to determine the most feasible and sensitive 3T MRI approach for cortical lesion detection by comparing IR-SWIET, T_1_w MP2RAGE, T_2_w fluid-attenuated inversion recovery (FLAIR), and T_1_/T_2_ ratio imaging in a multi-contrast reading paradigm.

Specifically, we evaluated whether the addition of IR-SWIET or T_1_/T_2_ improves cortical lesion detection compared with MP2RAGE and FLAIR alone, and how the number of IR-SWIET acquisitions (one, average of two or four repetitions, or a denoised single acquisition) affects cortical lesion detection.

## Materials and methods

### Data acquisition

Data collection was approved by the institutional review boards of the University of Vermont (UVM) and the National Institutes of Health (NIH), and informed consent was obtained from all participants. 71 pwMS underwent 3T brain MRI at UVM as a part of an observational study.

Scans for 11 participants were excluded due to artifacts. All participants underwent clinical testing including Expanded Disability Status Scale (EDSS), Symbol Digit Modalities Test (SDMT), timed 25-foot walk (T25FW), and 9-hole peg test (9-HPT). In addition, data from 10 pwMS who were enrolled in an observational study at the NIH with prior 3T brain MRI including IR-SWIET as well as 7T brain MRI were analyzed.^9^

### MRI acquisition and image processing

Brain MRIs at UVM were performed on a 3T Philips Achieva scanner (Philips Healthcare, Eindhoven, the Netherlands) equipped with a 32-channel head coil. The imaging protocol included IR-SWIET (acquired four times per session, acquired at 0.8 mm × 0.8 mm × 0.64 mm, reconstructed at 0.64 mm^3^ resolution), T_1_w MP2RAGE (1 mm^3^), and T_2_w FLAIR (0.89 × 0.89 × 1 mm). The four IR-SWIET acquisitions were rigidly co-registered (Advanced Normalization Tools [ANTs], version 2.5.3). Average of two (×2) and median of four (×4) IR-SWIET images were generated. A single acquisition was denoised (×1DN) using ANTs Denoise (patch radius = 1, search radius = 2, noise variance σ = 0.010); these conservative parameters were chosen to balance noise reduction with preservation of anatomical and pathological detail.^14^ The first acquisition was always used for IR-SWIET×1 and IR-SWIET×1DN, and the first two acquisitions were always used for IR-SWIET×2. FLAIR images were upsampled to 0.8 mm^3^, and all IR-SWIET images and MP2RAGE were rigidly registered (ANTs) to the upsampled FLAIR, using a mutual information cost function and linear interpolation. T_1_/T_2_ ratio images were generated by dividing the T_1_w MP2RAGE uniform denoised images (generated on the scanner) by the T_2_w FLAIR images. White matter lesions were segmented on FLAIR images using FLAMeS, followed by manual adjustment.^15^

For the NIH cohort, which was previously used to determine the sensitivity of cortical lesion detection using IR-SWIET, IR-SWIET (acquired four times) and FLAIR were obtained on a 3T Philips Achieva scanner and MP2RAGE on a 3T Siemens Magnetom Skyra scanner, each with a 32-channel head coil. 7T brain MRI included MP2RAGE (0.5 mm^3^, median of four acquisitions) and T_2_*-weighted gradient-recalled echo (GRE, 0.5 mm^3^) on a Siemens whole-body research system. Image processing details have been previously described.^9^

Detailed acquisition parameters for both cohorts are provided in Supplementary Table 1.

### Cortical lesion detection comparison (UVM cohort)

20 individuals from the UVM cohort were selected for cortical lesion detection comparison based on the availability of high-quality images and the presence of at least one visible cortical lesion identified on initial screening by an expert rater (ESB) with over eight years of cortical lesion segmentation experience. Participant demographics are summarized in Table 1.

**Table 1.** University of Vermont cohort demographic and clinical characteristics.

|  |  |
| --- | --- |
| **Female (n, %)** | 16 (80%) |
| **Age (years)<sup>1</sup>** | 47 ± 8 |
| **Clinical subtype (n, %)** |  |
| **Relapsing-remitting** | 17 (85%) |
| **Primary progressive** | 1 (5%) |
| **Secondary progressive** | 2 (10%) |
| **Years since diagnosis<sup>1</sup>** | 12 ± 8 |
| **Disease modifying therapy (n, %)** |  |
| **High efficacy** | 11 (55%) |
| **Moderate efficacy** | 1 (5%) |
| **Low efficacy** | 2 (10%) |
| **None** | 6 (30%) |
| **Expanded disability status scale<sup>2</sup>** | 2.5 (2.5) [1.5–7.0] |
| **9-hole peg test (dominant hand) (seconds)<sup>1</sup>** | 21.0 ± 5.1 |
| **Timed 25-foot walk (seconds)<sup>1</sup>** | 4.4 ± 0.9 |
| **Symbol Digit Modalities Test<sup>1</sup>** | 58 ± 8 |
| **T<sub>2</sub> weighted lesion volume (mm<sup>3</sup>)<sup>2</sup>** | 7071 (10124) [228–55362] |

Disease modifying therapy efficacy classifications were defined as: high efficacy (ocrelizumab, ofatumumab, cladribine); moderate efficacy (dimethyl fumarate, fingolimod, teriflunomide); low efficacy (glatiramer acetate, peginterferon beta-1a, interferon beta-1a).
<sup>1</sup>mean ± standard deviation
<sup>2</sup>median (interquartile range) [min – max]

For each of the 20 participants, cortical lesions were initially identified and segmented independently on five sets of images:

1. MP2RAGE + FLAIR
2. MP2RAGE + FLAIR + T_1_/T_2_ ratio images
3. MP2RAGE + FLAIR + IR-SWIET×1
4. MP2RAGE + FLAIR + IR-SWIET×2
5. MP2RAGE + FLAIR + IR-SWIET×4

Images for sets 1-5 above were divided into five reading sets, with each subject appearing once per set and equal representation of each of the first five multi-contrast image types within each set. Images were anonymized with respect to subject identity, demographic and clinical data, and IR-SWIET acquisition number. Subsequently, a sixth multi-contrast read:

1. MP2RAGE + FLAIR + IR-SWIET×1DN

was performed by the same two raters, with scans from each participant presented in a newly randomized order.

Cortical lesions were manually segmented using ITK-SNAP version 4.2.0. Images were viewed in axial, sagittal, and coronal planes simultaneously with linear interpolation enabled. Two raters (ES and KO) identified and segmented cortical lesions independently in the same order, with at least one week between each reading set. Criteria for cortical lesions included: (1) hypointense signal on MP2RAGE or T_1_/T_2_ and/or (2) hyperintensity on FLAIR and/or IR-SWIET; and (3) presence on two consecutive slices in at least one plane. Cortical lesions were categorized as leukocortical: involving both cortex and white matter but not extending to the pial surface, intracortical: confined to cortex but not extending to the pial surface; subpial: confined to cortex and touching the pial surface; and subpial with white matter (WM) involvement: extending from pial surface into white matter. Additional labels were used where definitive classification was not possible: juxtacortical vs leukocortical for lesions primarily located within the white matter but with possible cortical involvement; and possible subpial, possible intracortical, and possible leukocortical lesions. These additional lesion types were included in retrospective review analyses but not in primary lesion counts. A third rater (ESB) adjudicated all lesions identified only by one rater or for which there was disagreement on subtype.

### 7T validation of denoised IR-SWIET

To determine the sensitivity of IR-SWIET×1DN for cortical lesions, two raters independently identified cortical lesions on a combination of IR-SWIET×1DN, MP2RAGE, and FLAIR in 10 3T scans from participants with previous 7T-based as well as 3T-IR-SWIET-based cortical lesion segmentations (NIH cohort), using the same lesion criteria and rating protocol. Cortical lesion masks were compared to a reference standard mask previously generated using a combination of 7T T_1_w MP2RAGE×4 and T_2_*w GRE and to masks previously generated using 3T MP2RAGE, FLAIR, and IR-SWIET×2.^9, 16^

### Statistical analysis

Cortical lesion counts were compared across the six multi-contrast reads using the Friedman test, with p-values obtained by within-subject permutation (10,000 resamples) to accommodate the large number of tied values. Post-hoc pairwise comparisons used the Wilcoxon signed-rank test, with midranks for ties and zero differences discarded before ranking; exact two-sided p values were computed, with Benjamini–Hochberg false-discovery rate (FDR) correction within each of two pre-specified comparison families (Supplementary Table 2). Cortical lesion subtype-specific analyses used the same approach. The proportion of individuals with at least one detectable cortical or subpial lesion was compared across image sets using Cochran’s Q test, with pairwise comparisons performed using McNemar’s exact test with false-discovery rate (FDR) correction. Interrater reliability was assessed using intraclass correlation coefficient (ICC) using a two-way random effects model for total lesion counts and for each cortical lesion subtype per multi-contrast read, and 95% confidence intervals were computed by subject-level bootstrap comparison approach (5000 iterations, resampling with replacement) and reported descriptively. Partial Spearman’s rank correlations (ρ), adjusting for age, were calculated between cortical lesion counts and clinical measures for each multi-contrast read, and between white matter lesion volume and clinical measures. Age alone was included as a covariate given the sample size, and a nominal significance threshold of p < 0.05 (two-tailed) was used with no adjustment for multiple comparisons due to the exploratory nature of these analyses. In the 7T cohort, true positive counts and per-participant sensitivity were compared between IR-SWIET×1DN and IR-SWIET×2 using the Wilcoxon signed-rank test with the same tie handling and exact p values.

All statistical analyses were performed using Python 3.11.8.

## Results

### IR-SWIET increases detection of subpial cortical lesions, with similar performance of IR-SWIET×2, IR-SWIET×4, and IR-SWIET×1DN

Cortical lesion number was compared across six image sets in 20 individuals with MS (Table 2). Total cortical lesion number differed across image sets (Friedman χ² = 18.15, p = 0.002). Median cortical lesion number was similar in most pairwise comparisons, with the exception of higher cortical lesion number with IR-SWIET×2 (median 15, IQR 14; p = 0.011, FDR p = 0.042) and IR-SWIET×4 (median 16, IQR 14; p = 0.017, FDR p = 0.042) and lower with T_1_/T_2_ (median 8, IQR 11; p = 0.030, FDR p = 0.049) vs MP2RAGE + FLAIR alone (median 10, IQR 13).

**Table 2.** Cortical lesion counts on different image sets.

|  | Total cortical lesions |  | Leukocortical lesions |  | Intracortical lesions |  | Subpial lesions |  | Subpial with white matter involvement |  |
| --- | --- | --- | --- | --- | --- | --- | --- | --- | --- | --- |
| Image set<sup>1</sup> | Total | Median (IQR) [range] | Total | Median (IQR) [range] | Total | Median (IQR) [range] | Total | Median (IQR) [range] | Total | Median (IQR) [range] |
| MP2RAGE + FLAIR | 261 | 10 (13) [1–42] | 180 | 8 (9) [0–18] | 7 | 0 (0) [0–2] | 44 | 0 (2) [0–15] | 30 | 0 (3) [0–9] |
| T<sub>1</sub>/T<sub>2</sub> | 202 | 8 (11) [1–22] | 162 | 6 (11) [0–21] | 5 | 0 (0) [0–1] | 12 | 0 (1) [0–4] | 23 | 0 (2) [0–4] |
| IR-SWIET×1 | 261 | 12 (15) [0–27] | 172 | 8 (8) [0–20] | 2 | 0 (0) [0–1] | 57 | 1 (6) [0–10] | 30 | 0 (3) [0–5] |
| IR-SWIET×2 | 309 | 15 (14) [1–46] | 171 | 7 (10) [0–18] | 2 | 0 (0) [0–1] | 96 | 5 (5) [0–23] | 40 | 1 (2) [0–12] |
| IR-SWIET×4 | 318 | 16 (14) [0–47] | 169 | 8 (10) [0–20] | 6 | 0 (1) [0–1] | 103 | 4 (6) [0–27] | 40 | 1 (4) [0–8] |
| IR-SWIET×1DN | 289 | 14 (12) [0–47] | 151 | 6 (10) [0–21] | 0 | 0 (0) [0–0] | 88 | 4 (6) [0–22] | 50 | 1 (4) [0–11] |
| *Friedman test* | $\chi^2=18.15$, p=0.002 | | $\chi^2=3.23$, p=0.68 | | $\chi^2=12.41$, p=0.02 | | $\chi^2=42.00$, p<0.001 | | $\chi^2=25.12$, p<0.001 | |

<sup>1</sup>All image sets included MP2RAGE and FLAIR.
IR-SWIET: inversion recovery susceptibility weighted imaging with enhanced T<sub>2</sub> weighting; MP2RAGE: magnetization prepared 2 rapid acquisition gradient echoes; FLAIR: fluid-attenuated inversion recovery; IQR: interquartile range; IR-SWIET×1: single IR-SWIET acquisition; IR-SWIET×2: average of two acquisitions; IR-SWIET×4: median of four acquisitions; IR-SWIET×1DN: denoised single acquisition.

Subpial cortical lesions often appeared more conspicuous on IR-SWIET compared to other image types (Figure 1). Subpial lesion detection differed significantly across image sets (Friedman χ² = 42.00, p < 0.001). More subpial lesions were detected with IR-SWIET×2 (median 5, IQR 5; p = 0.002, FDR p = 0.008), IR-SWIET×4 (median 4, IQR 6; p = 0.003, FDR p = 0.008), and IR-SWIET×1DN (median 4, IQR 6; p = 0.005, FDR p = 0.008) compared to MP2RAGE + FLAIR alone (median 0, IQR 2). There was no increase in subpial lesion detection with IR-SWIET×1 (median 1, IQR 6, p = 0.42) (Table 2, Supplementary Table 2). There was no difference in subpial lesion detection using T_1_/T_2_ vs MP2RAGE + FLAIR alone (median 0, IQR 1, p = 0.056; FDR p = 0.07). Across IR-SWIET image sets, more subpial lesions were detected with IR-SWIET×2 (p = 0.03, FDR p = 0.03), ×4 (p = 0.01, FDR p = 0.03), and ×1DN (p = 0.02, FDR p = 0.03) than with IR-SWIET×1. The number of subpial lesions detected using IR-SWIET×1DN did not differ significantly from IR-SWIET×2 or ×4 (p = 0.41 for both).

**Figure 1.**
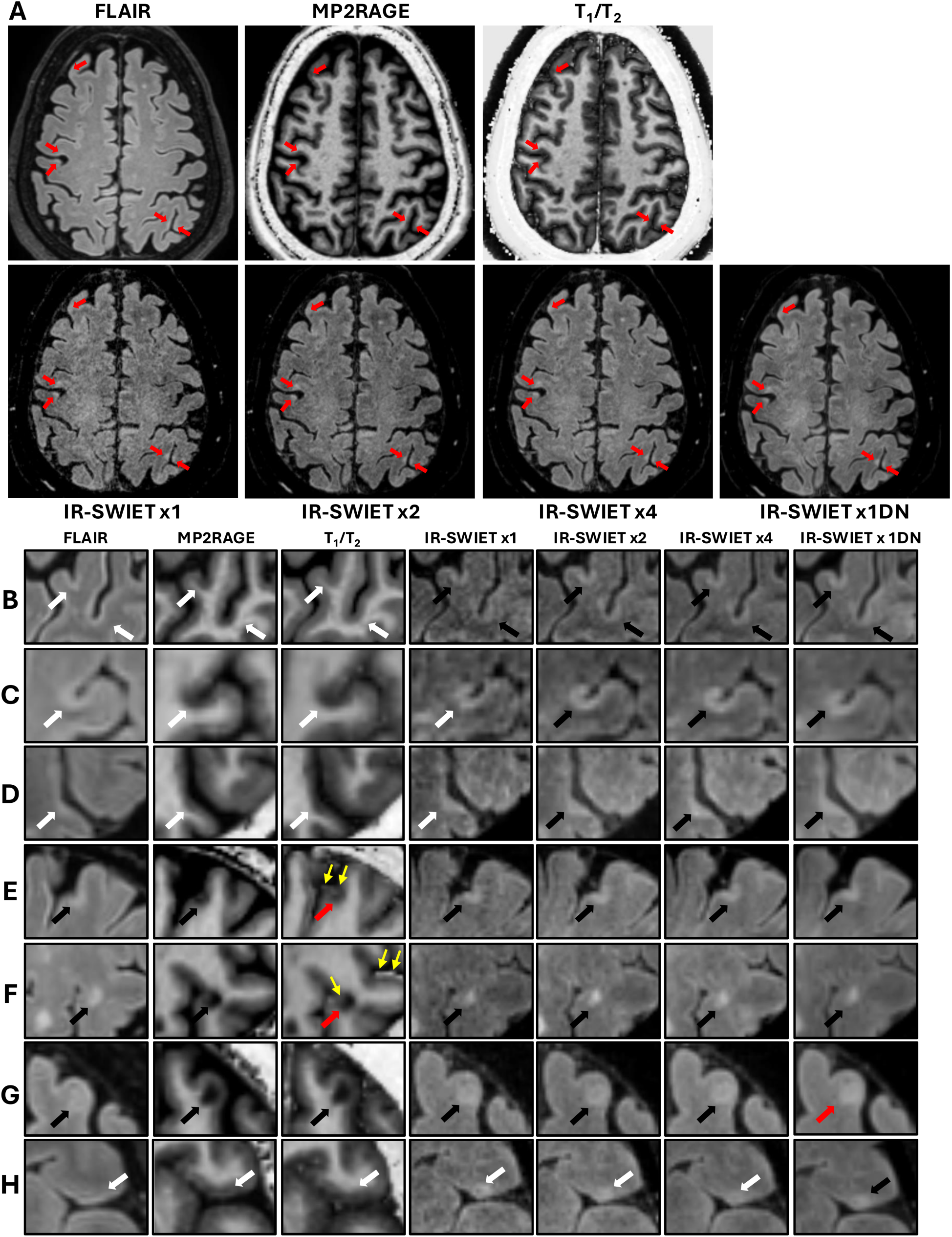
Cortical lesions are more conspicuous on IR-SWIET. (A) Axial slices of the image types used for cortical lesion identification: T_2_ weighted (w) fluid-attenuated inversion recovery (FLAIR), T_1_w magnetization-prepared 2 rapid acquisition gradient echoes (MP2RAGE), the ratio of MP2RAGE to FLAIR images (T_1_/T_2_), and Inversion Recovery Susceptibility Weighted Imaging with Enhanced T_2_ weighting (IR-SWIET). IR-SWIET was acquired four times per session and is shown as a single acquisition (×1), the average of two acquisitions (×2), the median of four acquisitions (×4), and a denoised single acquisition (×1DN). Red arrows indicate subpial lesions. (B–H) Magnified images of individual cortical lesions; black arrows indicate detected lesions; white, not detected; red, detected but misclassified; yellow, artifact. (B) Two subpial lesions identified on all IR-SWIET image types, with corresponding signal change visible on FLAIR and MP2RAGE in retrospect. (C-D) A subpial lesion identified on IR-SWIET×2, ×4, and ×1DN but not detected on FLAIR+MP2RAGE, T_1_/T_2_, or IR-SWIET×1. (E) A subpial lesion identified on FLAIR+MP2RAGE and all IR-SWIET image types but called possible subpial on T_1_/T_2_; yellow arrows indicate a division-related artifact on the T_1_/T_2_ image. (F) A leukocortical lesion identified on all image sets but classified as juxtacortical on T_1_/T_2_; yellow arrows indicate artifactual hyperintense signal at the pial surface. (G) A lesion identified as leukocortical on all image sets except IR-SWIET×1DN, where it was classified as subpial with white matter involvement, possibly related to smoothing of the lesion–cortex boundary by denoising. (H) A subpial lesion identified only on IR-SWIET×1DN but visible in retrospect on other image types.

Detection of subpial lesions with WM involvement also differed across image sets (Friedman χ² = 25.12, p < 0.001). More subpial lesions with WM involvement were detected with IR-SWIET×1DN compared to MP2RAGE + FLAIR (p = 0.001, FDR p = 0.005) and IR-SWIET×1 (p = 0.006, FDR p = 0.02). IR-SWIET×2 and IR-SWIET×4 did not differ significantly from MP2RAGE + FLAIR alone after FDR correction.

Leukocortical lesion detection did not differ significantly across image sets (Friedman χ² = 3.23, p = 0.68). Intracortical lesions were rare across all image sets (median 0 for all sequences); although there was a significant difference across image sets (Friedman χ² = 12.41, p = 0.02), no pairwise comparison survived FDR correction.

### Use of IR-SWIET increases the number of individuals with at least one detected subpial lesion

The proportion of individuals with at least one detected cortical lesion was high across all image sets (95–100%) and did not differ significantly across image sets (Cochran’s Q p = 0.63, Figure 2E). For subpial lesions, at least one subpial lesion was detected in 8/20 participants (40%) with MP2RAGE + FLAIR, vs 7/20 (35%) with T_1_/T_2_, 12/20 (60%) with IR-SWIET×1, 15/20 (75%) with IR-SWIET×2 and IR-SWIET×4, and 14/20 (70%) with IR-SWIET×1DN (Cochran’s Q, p < 0.001). More individuals had at least one detected subpial lesion with IR-SWIET×2 (p = 0.02, FDR p = 0.039) and IR-SWIET×4 (p = 0.02, FDR p = 0.039) compared with MP2RAGE +

**Figure 2.**
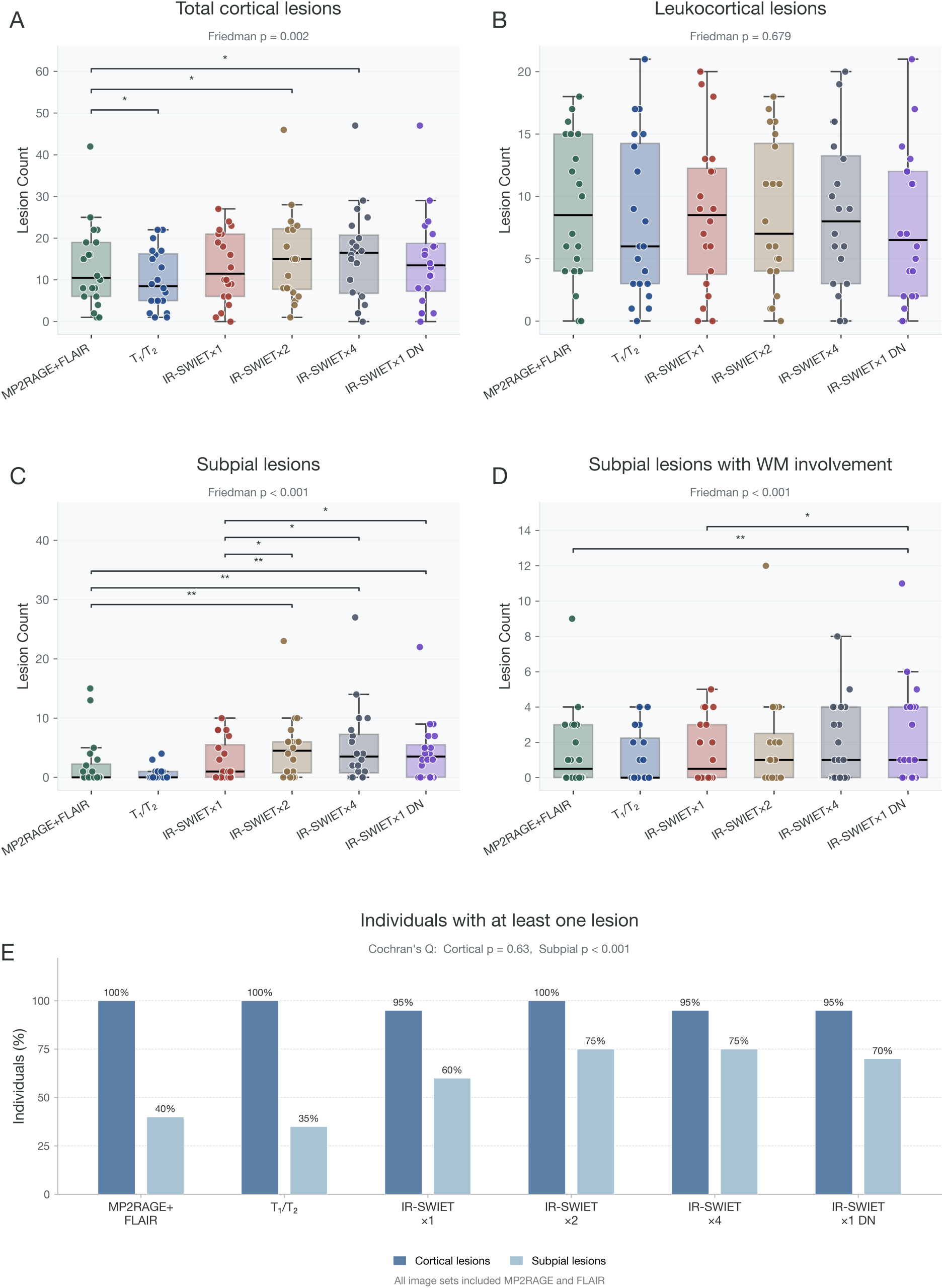
More subpial lesions are detected using IR-SWIET. (A–D) Boxplots of the number of cortical lesions identified on each image set in 20 participants. All image sets included fluid-attenuated inversion recovery (FLAIR) and magnetization-prepared 2 rapid acquisition gradient echoes (MP2RAGE). Each dot represents one individual; the center line indicates the median, the box the interquartile range, and the whiskers the highest and lowest values within 1.5 times the 1st and 3rd quartiles. The omnibus Friedman p value is shown above each panel. Significance brackets indicate pairwise comparisons against MP2RAGE + FLAIR (lower brackets) and against IR-SWIET×1 (upper brackets) (Wilcoxon signed-rank test with false discovery rate correction; * p < 0.05, ** p < 0.01; full results in Supplementary Table 2). Lesion counts were higher with the comparator image set in all significant comparisons except total cortical lesions on T_1_/T_2_, which were lower than on MP2RAGE + FLAIR. Intracortical lesions are not shown, as they were rare across all image sets (median 0). (E) The percentage of individuals with at least one detected cortical lesion (dark blue) was similar across image sets (95–100%; Cochran’s Q, p = 0.63), whereas the percentage with at least one subpial lesion (light blue) differed across image sets (Cochran’s Q, p < 0.001); pairwise McNemar comparisons are reported in the text and are not annotated here.

FLAIR; the difference in individuals with detected subpial lesions between IR-SWIET×1DN and MP2RAGE + FLAIR alone did not survive correction for multiple comparisons (p = 0.03, FDR p = 0.052).

### Interrater agreement for each image set

ICC estimates for total cortical, leukocortical, intracortical, and subpial lesions with white matter involvement showed no consistent pattern across image sets (Supplementary Table 3). ICC for subpial lesion counts was 0.66 (95% confidence interval [CI] 0.42 – 0.74) for IR-SWIET×4, 0.38 (CI 0.01 – 0.62) for IR-SWIET×2, 0.53 (CI 0.05 – 0.62) for IR-SWIET×1DN, 0.26 (CI -0.01 – 0.46) for IR-SWIET×1, 0.15 (CI 0.00 – 0.39) for T_1_/T_2_ and 0.29 (CI -0.09 – 0.79) for MP2RAGE + FLAIR (Supplementary Table 3).

The proportion of total cortical lesions identified independently by both raters was similar across image sets (53-65%). For subpial lesions, the proportion identified by both raters was 23% on MP2RAGE + FLAIR (10/44), 17% on T_1_/T_2_ (2/12), 37% on IR-SWIET×1 (21/57), 42% on IR-SWIET×2 (40/96), 50% on IR-SWIET×4 (51/103), and 56% on IR-SWIET×1DN (49/88).

### Retrospective review of cortical lesion appearance on different images

To better understand differences in lesion detection, we performed a retrospective review of specific lesion subsets.

#### Subpial lesions identified on IR-SWIET but not on MP2RAGE + FLAIR

Of the 178 lesions identified as subpial on at least one IR-SWIET-containing image set but not identified as subpial on MP2RAGE + FLAIR, 135 (76%) were not identified at all, 25 (14%) were identified as leukocortical, 16 (9%) were identified as possible subpial, and 2 (1%) were identified as intracortical. Of the 135 lesions not identified on MP2RAGE + FLAIR, for 90 (67%), there was subtle signal change on at least one sequence on retrospective review (Figure 1B-D) and no visible signal change in 31 (23%). 14 lesions (10%) were clearly visible and were considered missed lesions.

#### Subpial lesions identified only on IR-SWIET×1DN images

Of the 88 subpial lesions identified on IR-SWIET×1DN, 20 (23%) were not identified as subpial on any other image set. Of these, 11 (55%) were identified as possible subpial on at least one other image set, 7 (35%) were not detected on any other image set, and 2 (10%) were identified as subpial with WM involvement. Of the 7 subpial lesions not detected on any other image set, in retrospect, 4/7 (57%) were visible on IR-SWIET×4, with the lesion appearing more conspicuous on the denoised image in 2 lesions and equally visible in 2 lesions (Figure 1H). For the remaining 3 lesions (43%), there was subtle but not convincing signal abnormality on IR-SWIET×4, MP2RAGE, or FLAIR.

17 lesions were identified as subpial lesions with WM involvement on IR-SWIET×1DN but were not identified as such on any other image set. Of these, 11 (65%) were identified as leukocortical on at least one other image set, 2 (12%) were called subpial on one other image set (IR-SWIET×2), 1 (6%) was called possible subpial on one other image set (IR-SWIET×4), and 3 (18%) were not identified on any other image set. In some cases, the hyperintense subpial surface was confluent with the lesion on the denoised image, possibly leading to misclassification of leukocortical lesions as subpial lesions with WM involvement (Figure 1G).

#### Differences in lesion detection and classification on MP2RAGE + FLAIR vs T_1_/T_2_

The 65 lesions identified on MP2RAGE + FLAIR but not T_1_/T_2_ were clearly visible in retrospect (Supplementary Table 4). We observed that on the T_1_/T_2_ images there were artifacts related to imperfect registration and distortion differences that sometimes obscured lesions or made it difficult to determine if a lesion was juxtacortical vs cortical (Figure 1E–F).

### Associations between lesion counts and disability were similar across image sets

Associations between cortical lesion counts and clinical measures, adjusted for age, were broadly similar across image sets (Supplementary Figure 1). Higher total cortical lesion counts were associated with lower SDMT scores on all image sets except IR-SWIET×2 (ρ = -0.68 to -0.50; p < 0.05). Higher white matter lesion volume was also associated with lower SDMT scores (ρ = - 0.61, p = 0.004). Higher total cortical lesion counts detected on MP2RAGE + FLAIR were associated with longer 9-HPT times (ρ = 0.49, p = 0.03), but this association was not significant on any other image set. No significant associations with EDSS or T25FW were observed.

### Sensitivity of IR-SWIET×1DN is similar to IR-SWIET×2 when compared to 7T

To determine the sensitivity of cortical lesions detected on IR-SWIET×1DN, we performed a multi-contrast read using MP2RAGE, FLAIR, and IR-SWIET×1DN in a set of 10 scans from individuals with existing 3 and 7T-based cortical lesion segmentations. Cortical lesions identified on IR-SWIET×1DN were compared to previous 7T cortical lesion segmentations as well as to previous segmentations on MP2RAGE, FLAIR, and IR-SWIET×2.^9^ For total cortical lesions, IR-SWIET×1DN achieved a sensitivity of 34 ± 4% against the 7T reference, comparable to IR-SWIET×2 (sensitivity 40 ± 4%, p = 0.06). For subpial lesions, sensitivity with IR-SWIET×1DN was 25 ± 5%, similar to IR-SWIET×2 (sensitivity 30 ± 5%, p = 0.36, Table 3). Interrater agreement in this cohort was higher than in the UVM cohort (ICC 0.95, 95% CI 0.77 – 0.98 for total cortical lesions and 0.91, 0.49 – 0.97 for subpial lesions; Supplementary Table 5).

**Table 3.**
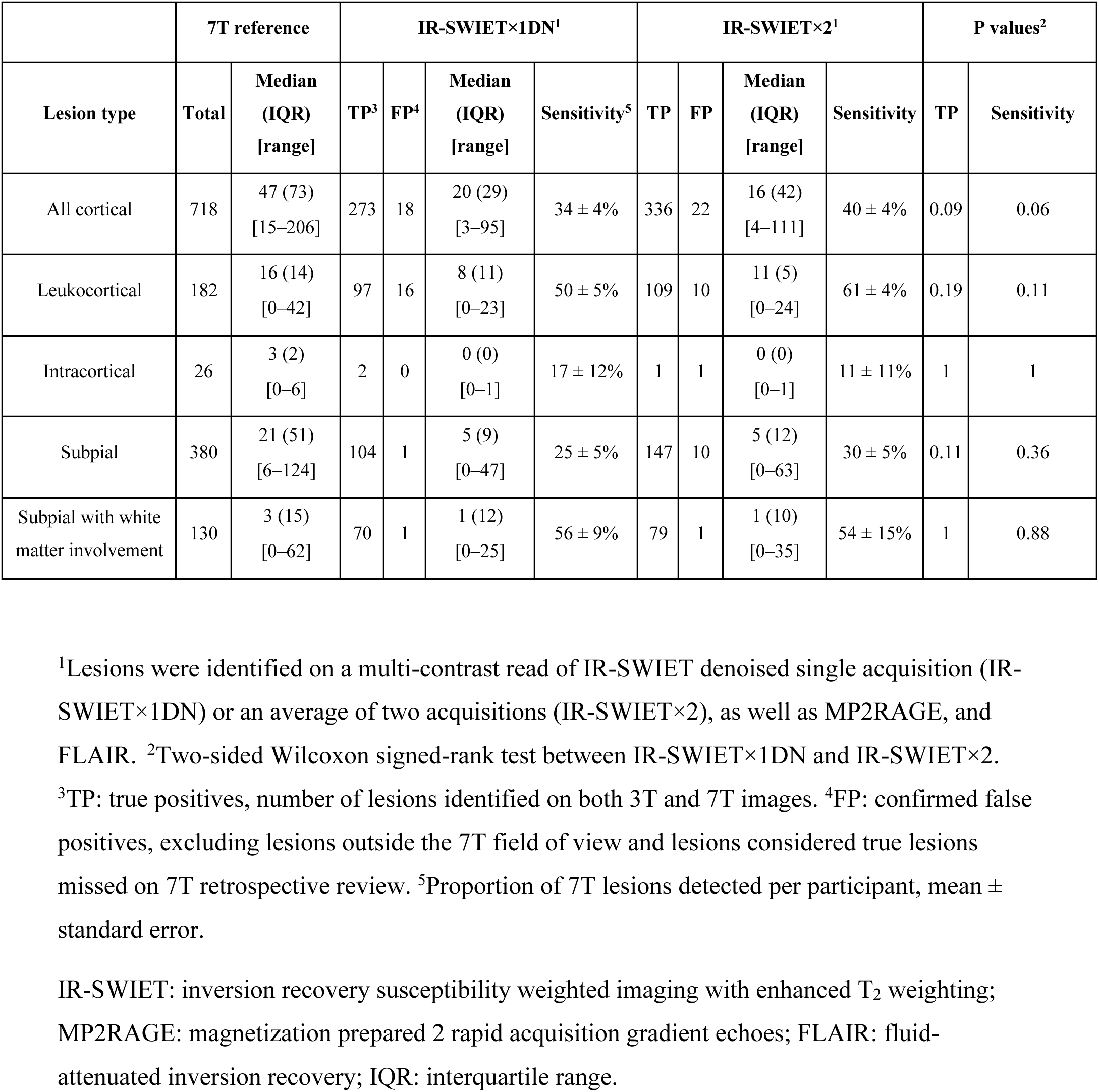
Comparison of cortical lesion detection using IR-SWIET×1DN with 7T reference standard.

|  | 7T reference |  | IR-SWIET×1DN<sup>1</sup> |  |  |  | IR-SWIET×2<sup>1</sup> |  |  |  | P values<sup>2</sup> |  |
| --- | --- | --- | --- | --- | --- | --- | --- | --- | --- | --- | --- | --- |
| Lesion type | Total | Median (IQR) [range] | TP<sup>3</sup> | FP<sup>4</sup> | Median (IQR) [range] | Sensitivity<sup>5</sup> | TP | FP | Median (IQR) [range] | Sensitivity | TP | Sensitivity |
| All cortical | 718 | 47 (73) [15–206] | 273 | 18 | 20 (29) [3–95] | 34 ± 4% | 336 | 22 | 16 (42) [4–111] | 40 ± 4% | 0.09 | 0.06 |
| Leukocortical | 182 | 16 (14) [0–42] | 97 | 16 | 8 (11) [0–23] | 50 ± 5% | 109 | 10 | 11 (5) [0–24] | 61 ± 4% | 0.19 | 0.11 |
| Intracortical | 26 | 3 (2) [0–6] | 2 | 0 | 0 (0) [0–1] | 17 ± 12% | 1 | 1 | 0 (0) [0–1] | 11 ± 11% | 1 | 1 |
| Subpial | 380 | 21 (51) [6–124] | 104 | 1 | 5 (9) [0–47] | 25 ± 5% | 147 | 10 | 5 (12) [0–63] | 30 ± 5% | 0.11 | 0.36 |
| Subpial with white matter involvement | 130 | 3 (15) [0–62] | 70 | 1 | 1 (12) [0–25] | 56 ± 9% | 79 | 1 | 1 (10) [0–35] | 54 ± 15% | 1 | 0.88 |

<sup>1</sup>Lesions were identified on a multi-contrast read of IR-SWIET denoised single acquisition (IR-SWIET×1DN) or an average of two acquisitions (IR-SWIET×2), as well as MP2RAGE, and FLAIR. <sup>2</sup>Two-sided Wilcoxon signed-rank test between IR-SWIET×1DN and IR-SWIET×2.
<sup>3</sup>TP: true positives, number of lesions identified on both 3T and 7T images. <sup>4</sup>FP: confirmed false positives, excluding lesions outside the 7T field of view and lesions considered true lesions missed on 7T retrospective review. <sup>5</sup>Proportion of 7T lesions detected per participant, mean ± standard error.
IR-SWIET: inversion recovery susceptibility weighted imaging with enhanced T<sub>2</sub> weighting; MP2RAGE: magnetization prepared 2 rapid acquisition gradient echoes; FLAIR: fluid-attenuated inversion recovery; IQR: interquartile range.

### Denoising IR-SWIET does not lead to an increase in false positive lesion detection

Of 327 cortical lesions identified on IR-SWIET×1DN, 54 (17%) were not originally identified on 7T images. On retrospective review of these lesions, 19 fell outside the 7T field of view and 4 could not be assessed owing to poor 7T image quality. Of the remaining 31, 13 were clearly visible on 7T images and were classified as missed (Figure 3B), and 18 (6% of all lesions identified) were considered false positives (16 leukocortical, one subpial, and one subpial with WM involvement, Figure 3D). Four of the leukocortical false positives appeared juxtacortical at 7T, and the other lesions did not have clear signal abnormality on 7T images. In comparison, 6% (22/358) lesions identified on IR-SWIET×2 were considered false positives, including 10 subpial lesions (Table 3).

**Figure 3.**
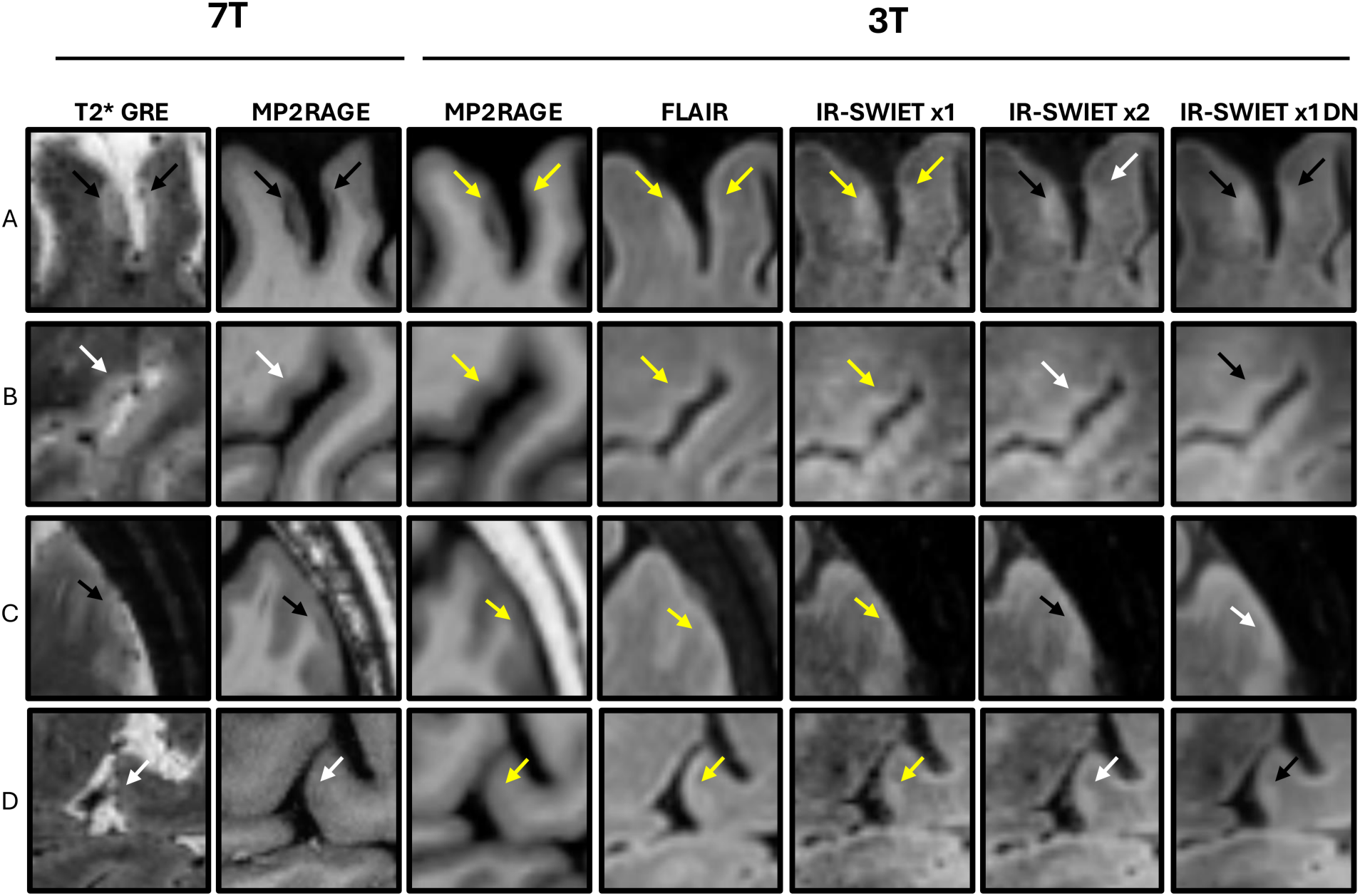
Cortical lesion detection on IR-SWIET×1DN, with comparison to IR-SWIET×2 and 7T imaging. Examples of cortical lesions on 7 tesla (T) and 3T images, including a single IR-SWIET acquisition (×1), an average of two acquisitions (×2), and a denoised single acquisition (×1DN). Black arrows indicate detected lesions; white, not detected. Cortical lesions were not identified on MP2RAGE and FLAIR alone or with IR-SWIET×1 here; yellow arrows therefore mark the lesion location on MP2RAGE, FLAIR, and IR-SWIET×1 without indicating whether the lesion was identified on those images. (A) Left: a subpial lesion identified on IR-SWIET×1DN, IR-SWIET×2, and at 7T. Right: a subpial lesion identified on IR-SWIET×1DN and at 7T, but not on IR-SWIET×2. (B) A subpial lesion identified on IR-SWIET×1DN that was not identified on 7T images but was visible on retrospective review. (C) A subpial lesion identified on IR-SWIET×2 and 7T images, but not on IR-SWIET×1DN, although it was visible on retrospective review. The superficial normal-appearing cortex appears more hyperintense on IR-SWIET×1DN than on IR-SWIET×2 and IR-SWIET×1, which may reduce lesion-to-cortex contrast and contribute to missed lesions on the denoised image. (D) A lesion identified on IR-SWIET×1DN but not on IR-SWIET×2 or on 7T images, considered a false positive on retrospective review, although subtle signal change was present on all IR-SWIET image types. T_2_* GRE: T_2_*-weighted gradient echo; MP2RAGE: magnetization-prepared 2 rapid acquisition gradient echoes; FLAIR: fluid-attenuated inversion recovery; IR-SWIET: Inversion Recovery Susceptibility Weighted Imaging with Enhanced T_2_ weighting.

### Cortical lesion subtyping accuracy is similar for IR-SWIET×1DN and ×2 vs 7T

We determined the proportion of cortical lesions assigned the same subtype based on 3T and 7T images. 79% of leukocortical lesions and 89% of subpial lesions were accurately classified on IR-SWIET×1DN. For subpial lesions with WM involvement, 47% were accurately classified on IR-SWIET×1DN while 41% were identified as subpial. Accuracy was similar for IR-SWIET×2 (see Supplementary Results).

## Discussion

Here we compared several 3T MRI approaches for cortical lesion detection. Our principal finding is that IR-SWIET significantly improves subpial cortical lesion detection compared to T_1_w MP2RAGE and T_2_w FLAIR imaging alone. In contrast, T_1_/T_2_ ratio imaging does not improve cortical lesion detection.

Consistent with our prior single-center study, IR-SWIET significantly improved subpial lesion detection.^9^ The increase in the proportion of individuals with at least one detectable subpial lesion from 40% with MP2RAGE and FLAIR to 70–75% with IR-SWIET is potentially clinically relevant for both diagnosis and prognosis.

Averaging two IR-SWIET acquisitions, which take ∼5 minutes each to acquire, and denoising a single acquisition both improved subpial lesion detection in comparison to using one IR-SWIET acquisition, with little additional gain from a median of four. There were no major differences in sensitivity, false positive rate, or subtyping accuracy between denoised IR-SWIET and averaged IR-SWIET, suggesting that denoising offers an efficient and accurate alternative. We did observe that denoising sometimes results in a more uniform, hyperintense subpial cortical signal, occasionally making it difficult to differentiate leukocortical from subpial lesions with WM involvement. More sophisticated deep-learning based denoising strategies, incorporating prior knowledge of cortical lesion appearance on averaged vs single IR-SWIET images, may in the future result in even better image quality and cortical lesion conspicuity.

Notably, adding T_1_/T_2_ reduced cortical lesion detection, despite previous findings of increased lesion-to-tissue contrast and lesion conspicuity with this method. This may have been due to artifacts on T_1_/T_2_ images, including hyperintense signal at the pial surface and changes in grey– white matter contrast, which may have obscured lesions or contributed to misclassification. Both artifacts likely arise from imperfect registration and geometric distortion differences between the two image types. The cortex-CSF artifact might be compounded by the use of FLAIR images as opposed to T_2_ weighted images to generate T_1_/T_2_; because FLAIR suppresses CSF signal, division by values approaching zero may contribute to artifact at the cortex-CSF boundary.^12,13^ Different registration approaches, intensity normalization before division, or acquisition of both sequences at matched resolution could potentially mitigate some of these issues.^13^

Interrater agreement for cortical lesions was modest for all image sets evaluated which is a recognized challenge for cortical lesion identification.^17^ Higher ICC values have been reported at 3T, 0.89 for total cortical lesions and 0.81 for subpial lesions with IR-SWIET×2, MP2RAGE and FLAIR and 0.67–0.87 for cortical lesions on phase-sensitive inversion recovery (PSIR) and FLAIR.^9,18,19^ However, ICC improved in the 7T validation with IR-SWIET×1DN, MP2RAGE and FLAIR to 0.95 for total cortical lesions and 0.91 for subpial lesions. Modest agreement observed in the UVM cohort could reflect rater inexperience and low overall lesion numbers in the UVM cohort rather than a limitation of IR-SWIET itself. Interrater limitations could be addressed with automated cortical lesion detection methods currently under development.^17^

Several limitations warrant acknowledgement. A 7T reference standard was not available for the UVM cohort, precluding estimation of sensitivity and evaluation of false positives. Participants were selected for the presence of at least one cortical lesion visible on MP2RAGE, FLAIR and IR-SWIET; this could have biased results in favor of IR-SWIET. Another limitation is that currently IR-SWIET requires an average or post processing denoising for maximal subpial lesion detection, limiting its current potential for clinical use. However, scanner manufacturers are now implementing deep-learning based image reconstructions that dramatically reduce noise for other sequences, and in the future these methods could potentially be applied to IR-SWIET.

Overall, IR-SWIET significantly improves subpial cortical lesion detection at 3T. Averaging two acquisitions provides substantial benefit over a single acquisition, and denoising a single acquisition is a time-saving alternative to averaging multiple acquisitions. Larger, multicenter studies will be needed to determine the generalizability of these findings and the utility of IR-SWIET-detected cortical lesions for MS diagnosis and prognosis.

## Data availability

The imaging and clinical data supporting the findings of this study are not publicly available because they contain information that could compromise participant privacy, but are available from the corresponding author on reasonable request and subject to institutional data-sharing agreements.

## Acknowledgements

We thank the participants for their time and contribution to this study, and the faculty and staff at the University of Vermont for their assistance with patient recruitment and study visits.

## Supplementary material

Supplementary material is available online.

