## Supplementary Material for "Increased subpial cortical lesion detection at 3 tesla using Inversion Recovery Susceptibility Weighted Imaging with Enhanced T2 Weighting (IR-SWIET)"

### Supplementary Results

<u>Cortical lesion subtyping accuracy of IR-SWIET×1DN vs 7T</u>

Of 97 lesions identified as leukocortical at 7T, 77 (79%) were also identified as leukocortical on IR-SWIET×1DN, 15 (15%) as subpial with WM involvement, and 5 (5%) as subpial. Of 104 lesions identified as subpial at 7T, 93 (89%) were also identified as subpial on IR-SWIET×1DN, 7 (7%) as subpial with WM involvement, and 4 (4%) as leukocortical. Of 70 lesions identified as subpial with WM involvement at 7T, 33 (47%) were also identified as subpial with WM involvement on IR-SWIET×1DN, 29 (41%) as subpial, and 8 (11%) as leukocortical. Agreement on IR-SWIET×2 was similar: of 109 lesions identified as leukocortical at 7T, 80 (73%) were also leukocortical on IR-SWIET×2, 20 (18%) subpial with WM involvement, and 9 (8%) subpial; of 147 identified as subpial at 7T, 122 (83%) were subpial on ×2, 18 (12%) subpial with WM involvement, and 7 (5%) leukocortical; of 79 identified as subpial with WM involvement at 7T, 34 (43%) were subpial with WM involvement on ×2, 24 (30%) leukocortical, and 21 (27%) subpial.

**Supplementary Table 1. MRI acquisition parameters.**

| | UVM cohort (3 tesla) | | | NIH cohort (3 tesla) | | | NIH cohort (7 tesla) | |
|---|---|---|---|---|---|---|---|---|
| **Parameter** | **IR-SWIET** | **MP2RAGE** | **FLAIR** | **IR-SWIET** | **MP2RAGE** | **FLAIR** | **MP2RAGE** | **$T_2$*w GRE** |
| Orientation | Sagittal | Sagittal | Sagittal | Sagittal | Sagittal | Sagittal | Axial | Axial |
| Acquisition | 3D | 3D | 3D | 3D | 3D | 3D | 3D | 2D |
| Voxel size (mm) | 0.8×0.8[1] | 1.0×1.0 | 0.89×0.89 | 0.8×0.8[1] | 1.0×1.0 | 1.0×1.0 | 0.5×0.5 | 0.5×0.5 |
| Slice thickness (mm) | 0.64 | 1.0 | 1.0 | 0.64 | 1.0 | 0.76 | 0.50 | 0.50 |
| FOV (mm) | 245×245×160 | 240×240×150 | 256×256×176 | 245×229×160 | 256×240×176 | 224×224×237 | 224×168×112 | 240×168×30 |
| TI (ms) | 2650 | 700/2500 | 1650 | 2650 | 700/2500 | 1650 | 800/2700 | N/A |
| TR/TE (ms) | 51/25 | 5000/2.9 | 4800/301 | 58/32 | 5000/2.9 | 4800/321 | 6000/5 | 4095/11.4–55.8 |
| Flip angle (°) | 12 | 4/5 | 90 | 12 | 4/5 | 90 | 4/5 | 70 |
| Scan time (min:sec) | 4:58[2] | 5:45 | 6:29 | 5:08[2] | 8:16 | 4:55 | 10:32[2] | 11:26[3] |

[1]Acquired at 0.8×0.8 mm, reconstructed to 0.64×0.64 mm. [2]Acquired four times. [3]Three partially overlapping volumes acquired for near full supratentorial coverage. IR-SWIET: inversion recovery susceptibility weighted imaging with enhanced $T_2$ weighting; MP2RAGE: magnetization prepared 2 rapid acquisition gradient echoes; FLAIR: fluid attenuated inversion recovery; GRE: gradient recalled echo; FOV: field of view (SI×AP×RL); TI: inversion time; TR: repetition time; TE: echo time; N/A: not applicable.

### Supplementary Table 2. Pairwise comparisons of cortical lesion detection

| | vs MP2RAGE + FLAIR | | | | | vs IR-SWIET×1 | | |
|---|---|---|---|---|---|---|---|---|
| **Lesion type** | **$T_1/T_2$** | **IR-SWIET×1** | **IR-SWIET×2** | **IR-SWIET×4** | **IR-SWIET×1DN** | **IR-SWIET×2** | **IR-SWIET×4** | **IR-SWIET×1DN** |
| Total cortical | **0.030 (0.049)** | 0.889 (0.889) | **0.011 (0.042)** | **0.017 (0.042)** | 0.308 (0.385) | 0.091 (0.136) | **0.041** (0.123) | 0.334 (0.334) |
| Leukocortical | 0.252 (0.545) | 0.570 (0.712) | 0.788 (0.788) | 0.327 (0.545) | 0.030 (0.151) | 0.843 (0.843) | 0.782 (0.843) | 0.455 (0.843) |
| Intracortical | 0.756 (0.945) | 0.245 (0.408) | 0.194 (0.408) | 1.000 (1.000) | 0.062 (0.312) | 1.000 (1.000) | 0.220 (0.661) | 0.494 (0.741) |
| Subpial | 0.056 (0.070) | 0.424 (0.424) | **0.002 (0.008)** | **0.003 (0.008)** | **0.005 (0.008)** | **0.034 (0.034)** | **0.011 (0.034)** | **0.024 (0.034)** |
| Subpial with white matter involvement | 0.434 (0.542) | 0.961 (0.961) | 0.161 (0.268) | 0.089 (0.222) | **0.001 (0.005)** | 0.273 (0.273) | 0.192 (0.273) | **0.006 (0.018)** |

Values shown are unadjusted p (False discovery rate [FDR]-corrected p) for pairwise Wilcoxon signed-rank tests (two-sided). FDR correction (Benjamini–Hochberg) applied separately per lesion type (5 comparisons in family 1, 3 comparisons in family 2). Bold lettering indicates significant after FDR correction ($p < 0.05$). For all significant comparisons, lesion number was lower with MP2RAGE + FLAIR (family 1, with the exception of $T_1/T_2$) or IR-SWIET×1 (family 2). Lesion count did not differ between IR-SWIET×2, IR-SWIET×4, and IR-SWIET×1DN. IR-SWIET: inversion recovery susceptibility weighted imaging with enhanced $T_2$ weighting; MP2RAGE: magnetization prepared 2 rapid acquisition gradient echoes; FLAIR: fluid attenuated inversion recovery. IR-SWIET×1: single IR-SWIET acquisition; IR-SWIET×2: average of 2 acquisitions; IR-SWIET×4: median of 4 acquisitions; IR-SWIET×1DN: denoised single acquisition.

**Supplementary Table 3. Intraclass correlation coefficients for cortical lesion counts across image sets.**

| | MP2RAGE + FLAIR | $T_1/T_2$ | IR-SWIET×1 | IR-SWIET×2 | IR-SWIET×4 | IR-SWIET×1DN |
|---|---|---|---|---|---|---|
| Total cortical | 0.55 (0.31, 0.76) | 0.54 (0.24, 0.83) | 0.35 (0.07, 0.57) | 0.58 (0.25, 0.70) | 0.59 (0.29, 0.78) | 0.50 (-0.01, 0.66) |
| Leukocortical | 0.68 (0.35, 0.86) | 0.54 (0.15, 0.79) | 0.50 (0.14, 0.79) | 0.67 (0.39, 0.87) | 0.46 (0.05, 0.75) | 0.52 (0.08, 0.82) |
| Intracortical | 0.00 (-0.43, 0.43) | -0.04 (-0.47, 0.39) | 0.00 (-0.43, 0.43) | -0.13 (-0.54, 0.32) | -0.04 (-0.47, 0.39) | -0.04 (-0.40, 0.40) |
| Subpial | 0.29 (-0.09, 0.79) | 0.15 (0.00, 0.39) | 0.26 (-0.01, 0.46) | 0.38 (0.01, 0.62) | 0.66 (0.42, 0.74) | 0.53 (0.05, 0.62) |
| Subpial with white matter involvement | 0.09 (-0.09, 0.68) | 0.53 (0.25, 0.81) | 0.45 (0.18, 0.72) | 0.42 (-0.03, 0.62) | 0.41 (0.13, 0.74) | 0.69 (0.16, 0.90) |

Values represent intraclass correlation coefficients (ICC), (95% confidence interval) using a two-way random effects model for absolute agreement, with confidence intervals computed by subject-level bootstrap (5,000 iterations, resampling with replacement). All reads included MP2RAGE and FLAIR. IR-SWIET: inversion recovery susceptibility weighted imaging with enhanced $T_2$ weighting; MP2RAGE: magnetization prepared 2 rapid acquisition gradient echoes; FLAIR: fluid attenuated inversion recovery. IR-SWIET×1: single IR-SWIET acquisition; IR-SWIET×2: average of 2 acquisitions; IR-SWIET×4: median of 4 acquisitions; IR-SWIET×1DN: denoised single acquisition.

**Supplementary Table 4. Identification and classification of cortical lesions on MP2RAGE + FLAIR vs $T_1/T_2$.**

| | **Concordant** | **Classified differently** | **Not detected** |
|---|---|---|---|
| **Detected on MP2RAGE + FLAIR** | | | |
| Subpial (n = 44) | 9 (20%) | 13 (30%)[a] | 22 (50%) |
| Leukocortical (n = 180) | 110 (61%) | 27 (15%)[b] | 43 (24%) |
| **Detected on $T_1/T_2$** | | | |
| Subpial (n = 12) | 9 (75%) | 1 (8%)[c] | 2 (17%) |
| Leukocortical (n = 162) | 110 (68%) | 26 (16%)[d] | 26 (16%) |

Values are number of lesions (% of the row total). Concordant: identified and classified as the same subtype on both image sets; Classified differently: detected on both image sets subtyped differently; Not detected: not identified on the other image set. MP2RAGE: magnetization prepared 2 rapid acquisition gradient echoes; FLAIR: fluid-attenuated inversion recovery

[a]4 possible subpial, 6 subpial with white matter (WM) involvement, 2 leukocortical, 1 intracortical.

[b]24 juxtacortical, 2 subpial with WM involvement, 1 intracortical.

[c]1 subpial with WM involvement

[d]16 juxtacortical, 5 subpial with WM involvement, 2 subpial, 2 possible subpial, 1 intracortical.

**Supplementary Table 5. Interrater agreement for cortical lesion counts in the 7T validation cohort with MP2RAGE, FLAIR and IR-SWIET×1DN.**

| Lesion type | ICC (2, 1) | 95% CI (bootstrap) |
|---|---|---|
| Total cortical | 0.95 | 0.77 – 0.98 |
| Leukocortical | 0.82 | 0.53 – 0.95 |
| Intracortical | -0.07 | -0.21 – 0.57 |
| Subpial | 0.91 | 0.49 – 0.97 |
| Subpial with white matter involvement | 0.93 | 0.54 – 0.99 |

Intraclass correlation coefficients (ICC) for single measures under a two-way random effects model for absolute agreement, for cortical lesion counts identified by two raters on a multicontrast read of IR-SWIET×1DN, MP2RAGE and FLAIR in the NIH cohort (n = 10). 95% confidence intervals were obtained by subject-level bootstrap (5,000 iterations, resampling participants with replacement). ICC for intracortical lesions is not interpretable given the small number of lesions identified (n = 2). IR-SWIET: inversion recovery susceptibility weighted imaging with enhanced $T_2$ weighting; ×1DN: denoised single acquisition; MP2RAGE: magnetization prepared 2 rapid acquisition gradient echoes; FLAIR: fluid attenuated inversion recovery.

### Supplementary Figures

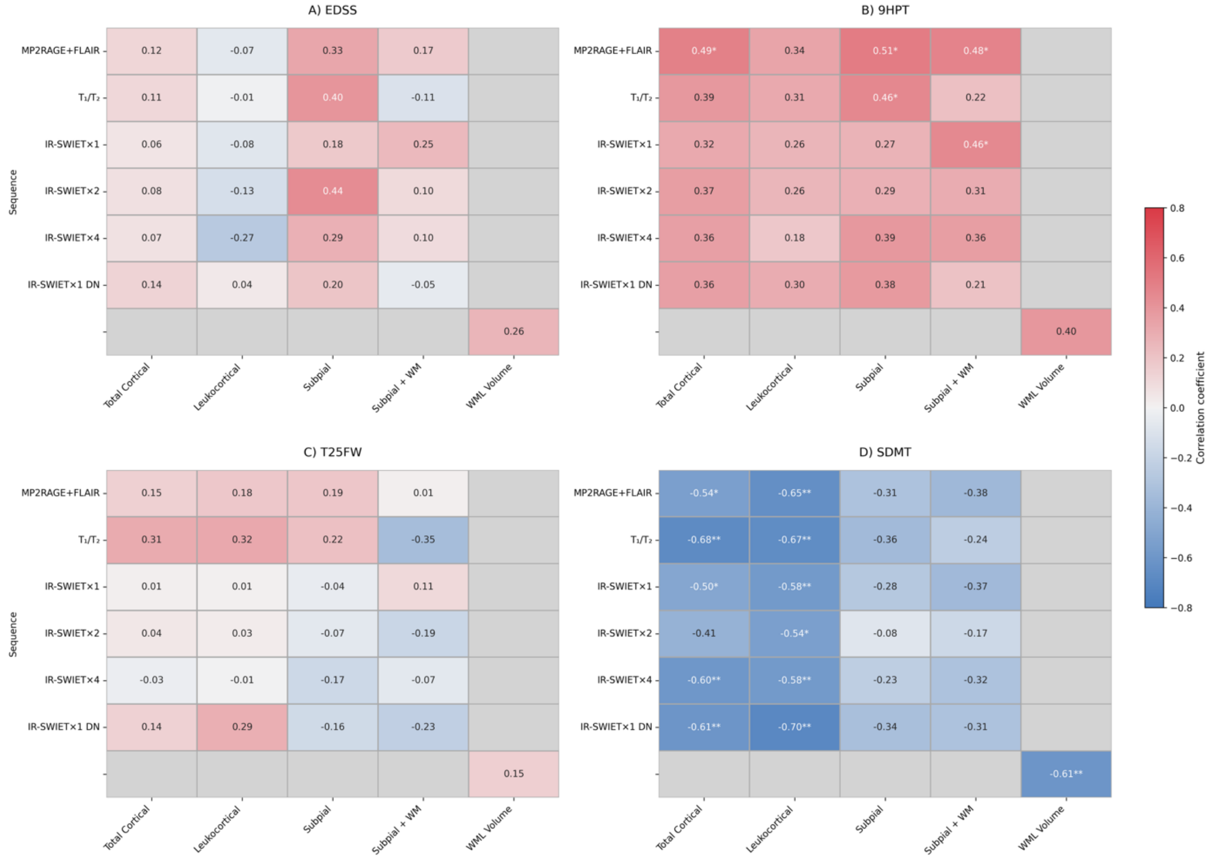


**Supplementary Figure 1. Associations between cortical lesion counts, white matter lesion volume, and clinical disability across image sets.** Heatmaps show partial Spearman correlation coefficients, adjusted for age, between lesion measures and clinical measures for each image set in the University of Vermont cohort (n = 20). Adjustment for age alone was used because of the small sample size. Lesion measures are total cortical, leukocortical, subpial, and subpial with white matter involvement lesion counts; intracortical lesions are not shown, as they were rare across all image sets (median 0). All image sets included magnetization-prepared 2 rapid acquisition gradient echoes (MP2RAGE) and fluid-attenuated inversion recovery (FLAIR). White matter lesion volume was measured on FLAIR and its association with each clinical measure, also adjusted for age, is therefore shown once, in the bottom-right cell of each panel. Grey cells indicate comparisons that are not applicable. Asterisks denote uncorrected significance (*$p < 0.05$, **$p < 0.01$); these analyses were not corrected for multiple comparisons and are exploratory. EDSS: expanded disability status scale; 9HPT: 9-hole peg test; T25FW:

timed 25-foot walk; SDMT: symbol digit modalities test; $T_1/T_2$: ratio of $T_1$ weighted to $T_2$ weighted images; IR-SWIET: inversion recovery susceptibility weighted imaging with enhanced $T_2$ weighting; ×1: single acquisition; ×2: average of 2 acquisitions; ×4: median of 4 acquisitions; ×1DN: single denoised acquisition.
